# Decoding the JAK-STAT axis in colorectal cancer with AI-HOPE-JAK-STAT: A conversational artificial intelligence approach to clinical-genomic integration

**DOI:** 10.1101/2025.06.08.25329216

**Authors:** Ei-Wen Yang, Brigette Waldrup, Enrique Velazquez-Villarreal

**Affiliations:** PolyAgent, San Francisco, CA; Department of Integrative Translational Sciences, Beckman Research Institute of City of Hope, Duarte, CA; City of Hope Comprehensive Cancer Center, Duarte, CA

**Author notes:** Corresponding Author –.

## Abstract

**Background/Objective:** The Janus kinase-signal transducer and activator of transcription (JAK-STAT) signaling pathway is a critical mediator of immune regulation, inflammation, and cancer progression. Although implicated in colorectal cancer (CRC) pathogenesis, its molecular heterogeneity and clinical significance remain insufficiently characterized—particularly within early-onset CRC (EOCRC) and across diverse treatment and demographic contexts. We present AI-HOPE-JAK-STAT, a novel conversational artificial intelligence platform built to enable real-time, natural language-driven exploration of JAK/STAT pathway alterations in CRC. The platform integrates clinical, genomic, and treatment data to support dynamic, hypothesis-generating analyses for precision oncology.

**Methods:** AI-HOPE-JAK-STAT combines large language models (LLMs), a natural language-to-code engine, and harmonized public CRC datasets from cBioPortal. Users define analytical queries in plain English, which are translated into executable code for cohort selection, survival analysis, odds ratio testing, and mutation profiling. To validate the platform, we replicated known associations involving JAK1, JAK3, and STAT3 mutations. Additional exploratory analyses examined age, treatment exposure, tumor stage, and anatomical site.

**Results:** The platform recapitulated established trends, including improved survival among EOCRC patients with JAK/STAT pathway alterations. In FOLFOX-treated CRC cohorts, JAK/STAT-altered tumors were associated with significantly enhanced overall survival (p < 0.0001). Stratification by age revealed survival advantages in younger (age <50) patients with JAK/STAT mutations (p = 0.0379). STAT5B mutations were enriched in colon adenocarcinoma and correlated with better outcomes (p = 0.0000). Conversely, JAK1 mutations in microsatellite-stable tumors did not affect survival, emphasizing the value of molecular context. Finally, JAK3-mutated tumors diagnosed at Stage I–III showed superior survival compared to Stage IV cases (p = 0.00001), reinforcing stage as a dominant clinical determinant.

**Conclusion:** AI-HOPE-JAK-STAT establishes a new standard for pathway-level interrogation in CRC by empowering users to generate and test clinically meaningful hypotheses without coding expertise. This system enhances access to precision oncology analyses and supports scalable, real-time discovery of survival trends, mutational associations, and treatment-response patterns across stratified patient cohorts.

## Introduction

Colorectal cancer (CRC) remains one of the most prevalent and lethal malignancies worldwide, ranking third in global cancer incidence and second in cancer-related deaths [1,3]. While overall CRC rates have stabilized or declined in older adults, the incidence of early-onset colorectal cancer (EOCRC)—defined as diagnosis before the age of 50— continues to rise across several populations with disproportionate health burdens in the United States [4–8]. This increase has been accompanied by high rates of EOCRC-related mortality [9–11].

EOCRC exhibits distinct clinical and molecular features, including a higher frequency of microsatellite instability (MSI), increased tumor mutational burden, immune checkpoint activation, and unique epigenetic signatures [12–16]. These characteristics suggest alternative oncogenic mechanisms that may differ from those driving late-onset CRC (LOCRC), warranting further investigation into pathway-specific alterations across age and ancestry groups.

The Janus kinase-signal transducer and activator of transcription (JAK/STAT) pathway is a major signaling cascades involved in CRC pathogenesis. The JAK/STAT pathway remains underexplored in EOCRC. Aberrant JAK/STAT signaling has been associated with chronic inflammation, tumor immune evasion, and epithelial-mesenchymal transition (EMT), contributing to more aggressive tumor behavior [1, 17–22] and reduced treatment response in CRC [23–27]. STAT3 activation, in particular, has been linked to stromal invasion and adverse survival outcomes [28–30].

Recent studies investigating molecular alterations in JAK/STAT pathway genes among CRC patients have reported higher frequencies of JAK/STAT-related mutations in EOCRC cases among individuals [31–33] from populations with different ethnicity [1].

However, no significant differences in JAK/STAT pathway alterations were observed across age or ancestry groups. Survival analyses indicated that JAK/STAT alterations may still be associated with clinical outcomes, although findings were variable across subgroups. These observations highlight the need for tools that can enable detailed, flexible exploration of molecular data across clinical and genetic contexts.

Advances in artificial intelligence (AI), including the application of large language models (LLMs), are transforming the ability to extract insights from complex biomedical datasets [34–38]. Conversational AI systems can interpret natural language queries and execute analytical workflows in real time, making it easier for researchers and clinicians to interact with genomic and clinical data. Despite their promise, most existing platforms lack customization for pathway-level interrogation or integration with multiple data modalities.

To meet this need, we developed AI-HOPE-JAK-STAT, a conversational AI agent specifically designed to investigate JAK/STAT pathway alterations in CRC. The system enables integration of clinical, genomic, and transcriptomic data through natural language–driven analysis. In this study, we describe the development of AI-HOPE-JAK-STAT, demonstrate its analytical performance by replicating known trends in CRC cohorts, and explore its application in identifying clinically relevant patterns in EOCRC across population groups.

## Methods

### AI-HOPE-JAK-STAT Platform Design and Functionality

AI-HOPE-JAK-STAT is a custom-built conversational artificial intelligence platform designed to explore the clinical and genomic landscape of JAK/STAT pathway alterations in CRC. The system leverages a LLM-driven interface that translates plain language queries into executable bioinformatics commands. This architecture enables real-time integration and interrogation of multi-dimensional CRC datasets using a modular backend combining data parsing, statistical modeling, and dynamic visualization.

### Data Sources and Processing

The platform ingests harmonized CRC datasets derived from the publicly available repository cBioPortal. Data preprocessing involved normalization of sample identifiers, removal of incomplete clinical records, and mapping of genomic features to canonical gene symbols based on HGNC nomenclature. Genes involved in the JAK/STAT signaling pathway—such as JAK1, JAK2, JAK3, TYK2, STAT1, STAT3, STAT5A/B, and SOCS family members—were preselected for targeted analysis. Clinical annotations included tumor stage, anatomical location, treatment history, microsatellite instability (MSI) status, and self-reported ancestry.

### Natural Language Input and Query Interpretation

Users interact with the platform through an intuitive natural language interface. AI-HOPE-JAK-STAT processes these inputs using an LLM (LLaMA 3-based) to identify intent, extract parameters, and structure downstream analytic operations. Example queries include: “Compare mutation rates in STAT3 between patients under and over 50,” or “Show survival differences in JAK1-mutated tumors treated with chemotherapy.” Ambiguities in user input trigger clarification prompts to maintain analytic precision and reproducibility.

### Analytical Framework

Backend analytics for AI-HOPE-JAK-STAT are implemented in Python, leveraging libraries such as *pandas*, *lifelines*, and *SciPy* to execute a range of statistical and bioinformatics operations. The platform performs descriptive summaries to assess mutation frequencies and characterize cohort composition. For categorical comparisons, it employs chi-square or Fisher’s exact tests to evaluate statistical significance. Survival outcomes are analyzed using Kaplan-Meier estimates with log-rank testing, while multivariable survival modeling is conducted using Cox proportional hazards regression. The system also enables flexible subgroup stratification based on age group, self-reported ancestry, treatment exposure, and tumor location. Additionally, AI-HOPE-JAK-STAT supports co-occurrence and mutual exclusivity analyses to explore potential interactions between JAK/STAT pathway genes and other molecular drivers relevant to colorectal tumorigenesis.

### Application Validation

To evaluate AI-HOPE-JAK-STAT’s analytical robustness, we applied the platform to replicate findings from previous large-cohort studies investigating JAK/STAT and MAPK pathway alterations across EOCRC and LOCRC populations. Validation tasks included mutation frequency comparisons between different ethnicity EOCRC patients and assessment of survival impact among STAT3-mutated cases.

### Output Generation and Interpretation

Following analysis, the system generates comprehensive visual and tabular outputs. These include survival curves, mutation distribution plots, heatmaps, and odds ratio forest plots. Results are accompanied by auto-generated narrative summaries linking observed patterns to published biomedical literature through a retrieval-augmented generation (RAG) module. All outputs are exportable for downstream reporting or figure preparation.

### Usability and Comparative Evaluation

To assess platform usability, we benchmarked AI-HOPE-JAK-STAT against existing tools (e.g., cBioPortal, Xena Browser) across common analytic tasks such as stratified mutation analysis, co-alteration assessment, and treatment-outcome comparisons.

Metrics evaluated included task completion time, flexibility in cohort definition, and reproducibility of results. AI-HOPE-JAK-STAT demonstrated increased efficiency, especially in generating nested subgroup analyses and complex multi-parameter queries.

## Results

AI-HOPE-JAK-STAT enabled natural language-driven interrogation of the JAK/STAT signaling cascade across diverse CRC patient populations, integrating genomic alterations with clinical outcomes in real time. Through a series of validation and exploratory analyses, the platform demonstrated its ability to reproduce known associations, uncover novel patterns, and generate interpretable visual and statistical outputs that facilitate hypothesis generation and precision oncology discovery.

To assess ancestry-specific differences in JAK/STAT alterations, we first examined EOCRC patients of H/L descent. Among individuals under 50 years of age, those with JAK/STAT pathway mutations (n = 16) showed a trend toward improved overall survival compared to those without such alterations (n = 137), though the result did not meet conventional statistical significance (p = 0.0539; Fig. 2). A similar query conducted in EOCRC patients of Non-Hispanic White (NHW) ancestry yielded a significant survival advantage for the JAK/STAT-altered group (p = 0.0001; Fig. S1), reinforcing the potential prognostic value of pathway-specific mutations in defined populations.

**Figure 1.**
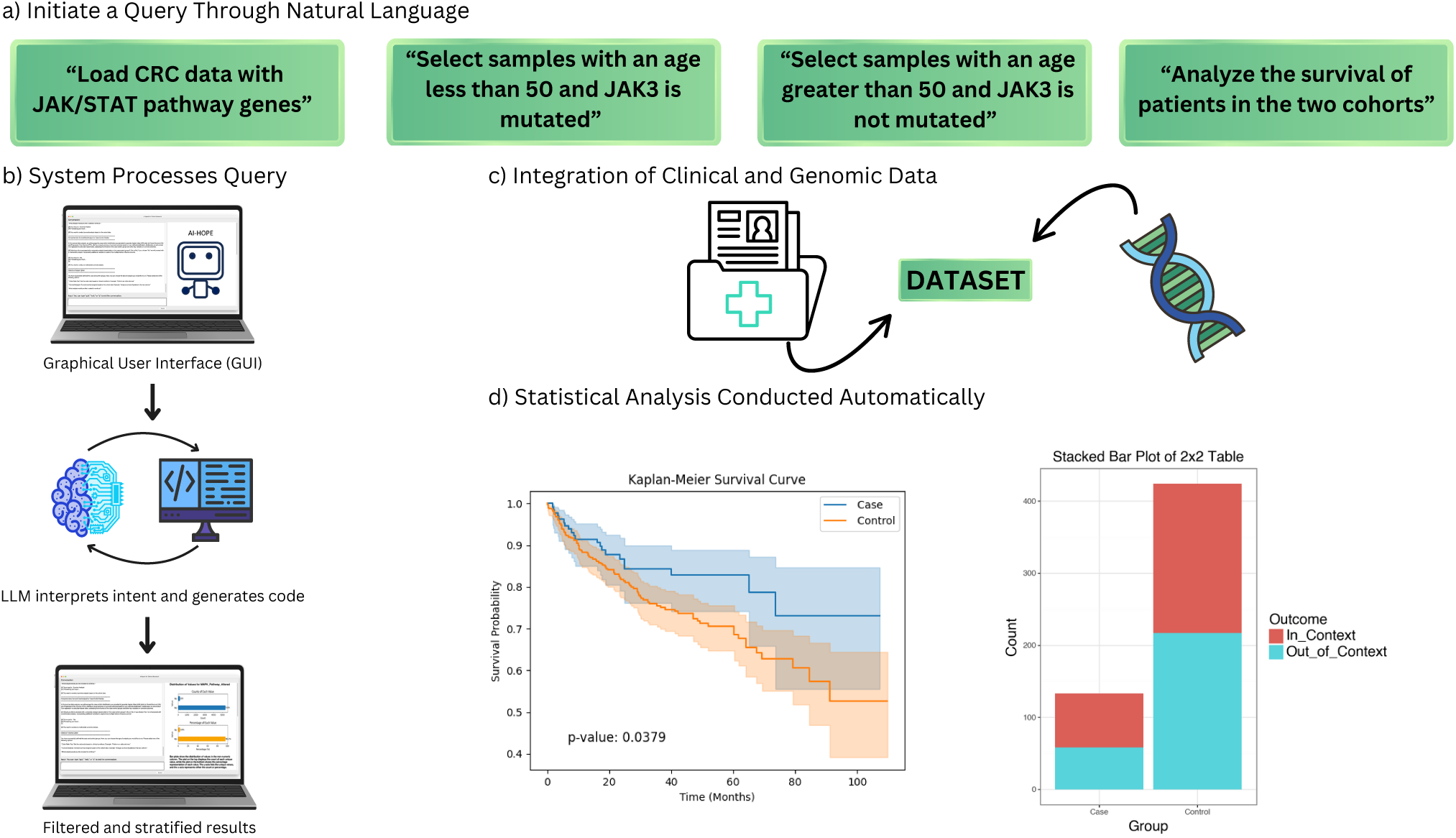
Workflow of AI-HOPE-JAK-STAT for clinical-genomic analysis of the JAK/STAT pathway in colorectal cancer. This figure illustrates the operational framework of AI-HOPE-JAK-STAT, a conversational artificial intelligence system designed to explore JAK/STAT signaling alterations in colorectal cancer (CRC) using natural language input. a) Users initiate analysis by entering plain language queries such as loading CRC datasets with JAK/STAT pathway genes, filtering by age or mutation status (e.g., JAK3), and requesting survival comparisons between defined cohorts. b) The system interprets the query through a graphical user interface (GUI) supported by a large language model (LLM), which parses the user’s intent, generates code, and executes the relevant operations. c) Clinical and genomic data are automatically retrieved and integrated from harmonized datasets. These include patient-level variables and mutation profiles relevant to the JAK/STAT signaling axis. d) Statistical analyses are then conducted automatically, producing visual outputs such as Kaplan-Meier survival curves and contingency plots. Results are stratified according to user-defined parameters and delivered alongside narrative interpretations, enabling streamlined investigation of JAK/STAT-driven tumor biology in CRC.

**Figure 2.**
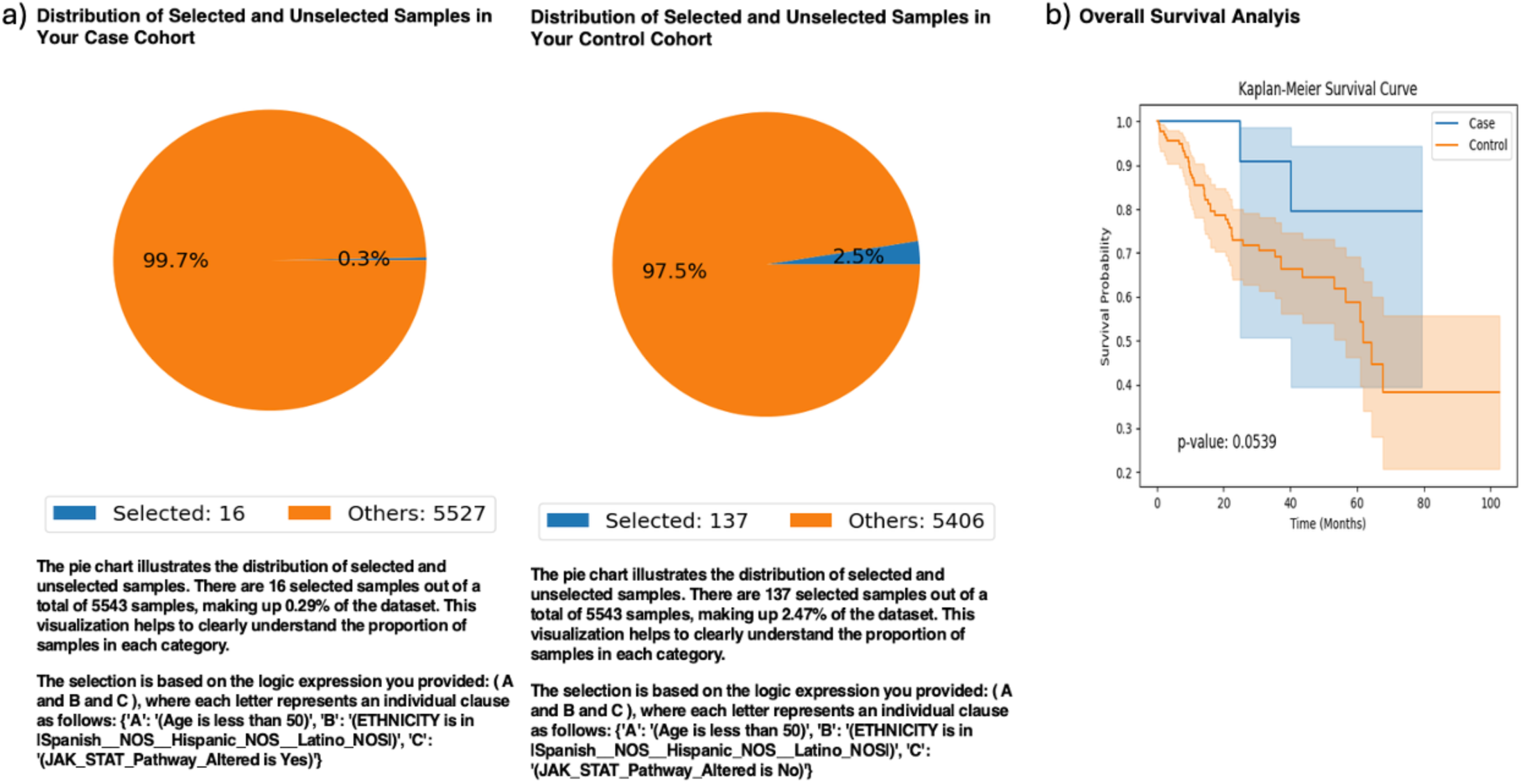
AI-HOPE-JAK-STAT analysis of early-onset colorectal cancer patients of ethnicity-specific descent stratified by JAK/STAT pathway alteration status. This figure presents the output of a natural language query executed via AI-HOPE-JAK-STAT to investigate the impact of JAK/STAT pathway alterations on overall survival in early-onset colorectal cancer (EOCRC) patients of Hispanic/Latino descent (H/L). The case cohort includes patients under age 50 with alterations in JAK/STAT pathway genes, while the control cohort includes similarly aged patients of the same ancestry group without such alterations. a) Pie charts display the proportional representation of selected samples in each cohort relative to the full dataset (n = 5543), highlighting the rarity of JAK/STAT pathway alterations in this subgroup. b) Kaplan-Meier survival analysis shows a trend toward improved survival in the JAK/STAT-altered cohort (blue) compared to the non-altered group (orange). Although the p-value of 0.0539 from the log-rank test does not reach conventional statistical significance (p < 0.05), the observed difference suggests a possible survival benefit that may warrant further validation in larger cohorts. Shaded confidence intervals illustrate variability in the estimates and highlight the importance of cautious interpretation.

Next, we investigated treatment context by focusing on CRC patients who received FOLFOX chemotherapy. Kaplan-Meier survival analysis revealed that individuals with JAK/STAT pathway alterations (n = 282) had significantly better survival compared to those without such mutations (n = 3,791) under the same treatment regimen (p < 0.0001; Fig. 3). These findings suggest that JAK/STAT alterations may be associated with enhanced treatment response in FOLFOX-exposed patients.

**Figure 3.**
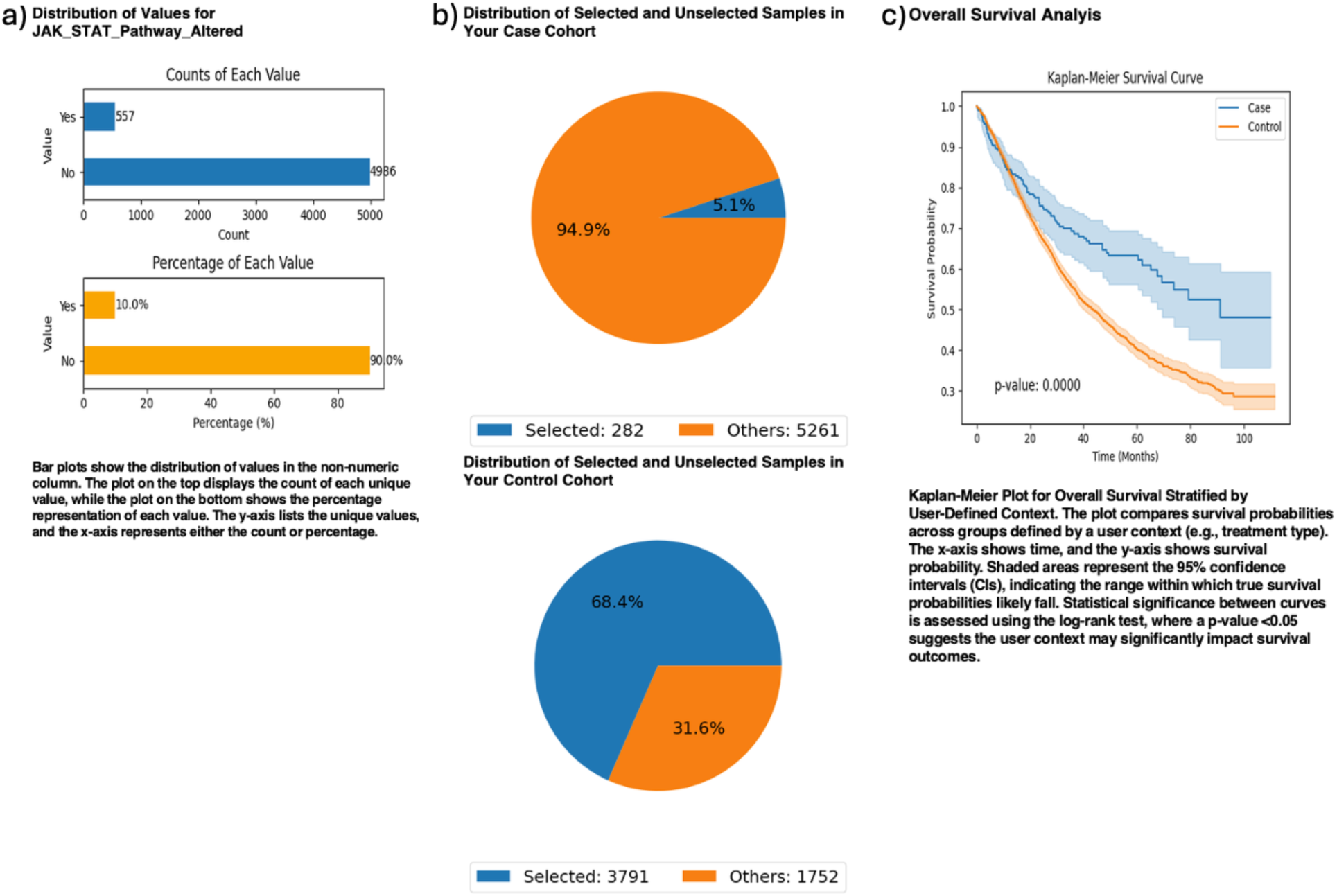
AI-HOPE-JAK-STAT analysis of colorectal cancer patients treated with FOLFOX stratified by JAK/STAT pathway alteration status. This figure presents the results of a natural language-driven analysis conducted through AI-HOPE-JAK-STAT, evaluating overall survival among colorectal cancer (CRC) patients treated with combination chemotherapy (Fluorouracil, Leucovorin, and Oxaliplatin—FOLFOX), stratified by JAK/STAT pathway alteration status. The case cohort includes patients with JAK/STAT alterations who received FOLFOX (n = 282), while the control cohort includes patients without such alterations who received the same regimen (n = 3,791). a) Bar plots summarize the distribution of JAK/STAT pathway alterations across the entire dataset (n = 5,543), showing that 10% of patients exhibit alterations in this pathway. The top chart displays absolute counts, while the bottom chart presents percentages. b) Pie charts illustrate the relative representation of selected patients in each cohort. The case cohort accounts for 5.1% of the dataset, while the control cohort constitutes 68.4%, reflecting the larger proportion of patients without JAK/STAT alterations receiving FOLFOX. c) Kaplan-Meier survival analysis reveals a statistically significant survival advantage in the JAK/STAT-altered cohort (blue) compared to the control group (orange), with a p-value < 0.0001. The clear separation between curves and narrow confidence intervals suggest a robust association between JAK/STAT pathway alterations and improved treatment outcomes among CRC patients undergoing FOLFOX chemotherapy.

To explore age-related effects, we stratified JAK/STAT-altered CRC patients by early-(<50 years; n = 133) versus late-onset (≥50 years; n = 424) status. Early-onset cases exhibited significantly improved survival (p = 0.0379), and odds ratio testing suggested modest enrichment of FOLFOX treatment in this younger group, although not statistically significant (OR = 1.356, p = 0.154; Fig. 4). These results highlight possible interactions between age, treatment exposure, and molecular alterations.

**Figure 4.**
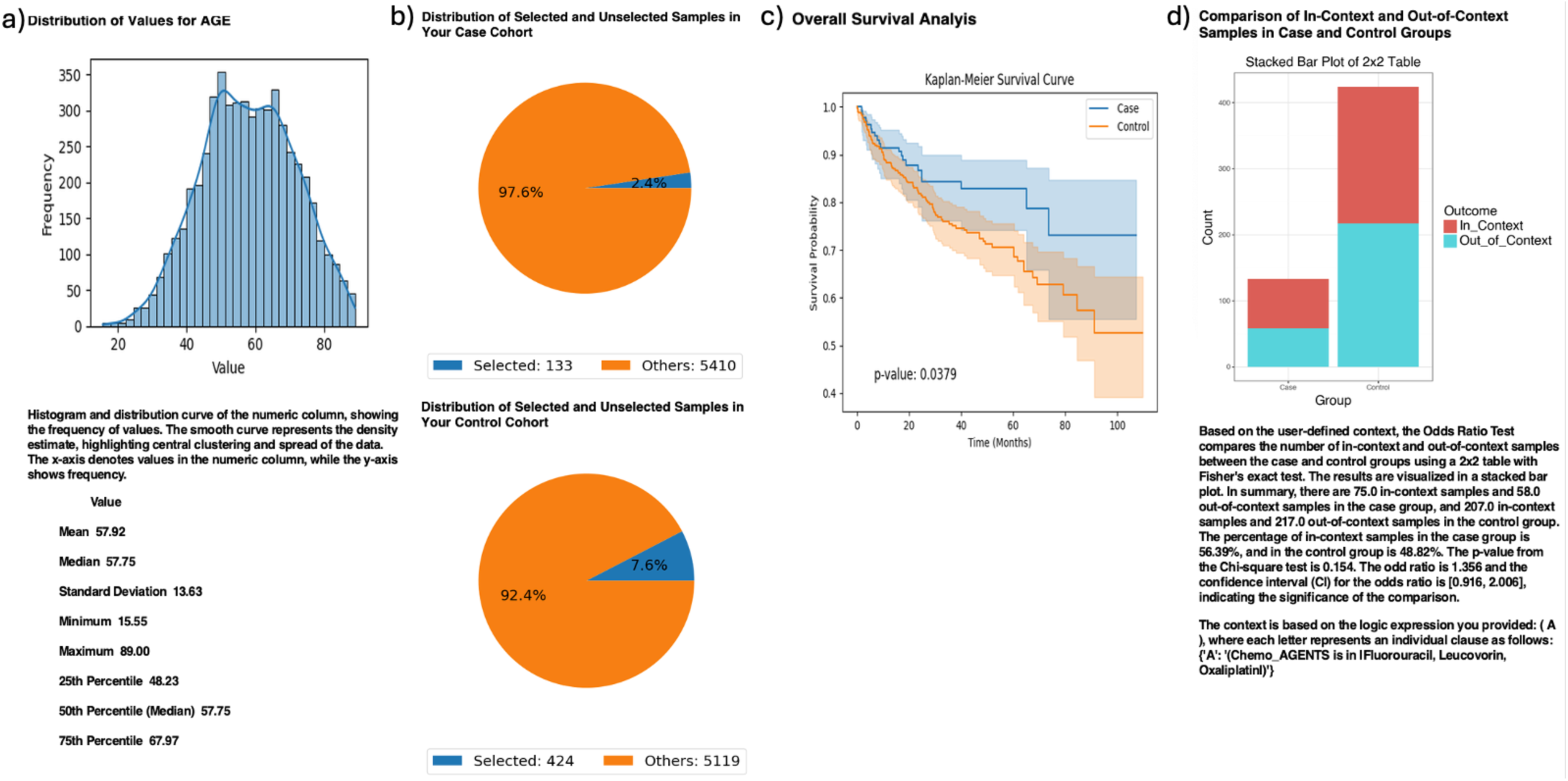
AI-HOPE-JAK-STAT analysis of age-stratified colorectal cancer patients with JAK/STAT pathway alterations treated with FOLFOX. This figure summarizes the results of a natural language query executed via AI-HOPE-JAK-STAT to compare survival outcomes and chemotherapy context between colorectal cancer (CRC) patients with JAK/STAT pathway alterations, stratified by age. The case cohort consists of patients younger than 50 years (n = 133), and the control cohort includes patients older than 50 (n = 424). All individuals received combination chemotherapy with Fluorouracil, Leucovorin, and Oxaliplatin (FOLFOX). a) A histogram displays the age distribution of the full dataset (n = 5,543), with a mean age of 57.92 years, confirming the rationale for early-vs. late-onset cohort definitions. b) Pie charts visualize the proportion of selected samples within each cohort relative to the entire population. The early-onset (case) group comprises 2.4% of samples, while the late-onset (control) group represents 7.6%. c) Kaplan-Meier survival analysis reveals a statistically significant survival difference between the two age groups, with the early-onset JAK/STAT-altered cohort (blue) demonstrating superior outcomes compared to the older group (orange) (p = 0.0379). d) A 2×2 odds ratio analysis was conducted to assess treatment context by comparing the number of in-context and out-of-context samples (based on receipt of FOLFOX chemotherapy) across cohorts. The stacked bar plot shows that 56.39% of early-onset cases and 48.82% of late-onset controls were in the user-defined treatment context. The resulting odds ratio was 1.356 (95% CI: 0.916–2.006, p = 0.154), suggesting a modest but non-significant enrichment of FOLFOX treatment in the early-onset group.

We then assessed the prognostic impact of JAK1 mutations in microsatellite stable (MSS) tumors. Among MSS CRC patients, those with JAK1 mutations (n = 44) showed no significant difference in overall survival relative to wild-type controls (n = 4,638; p = 0.5691; Fig. 5). Despite the absence of survival benefit, this analysis underscores the utility of AI-HOPE-JAK-STAT in isolating molecular contexts for precision stratification.

**Figure 5.**
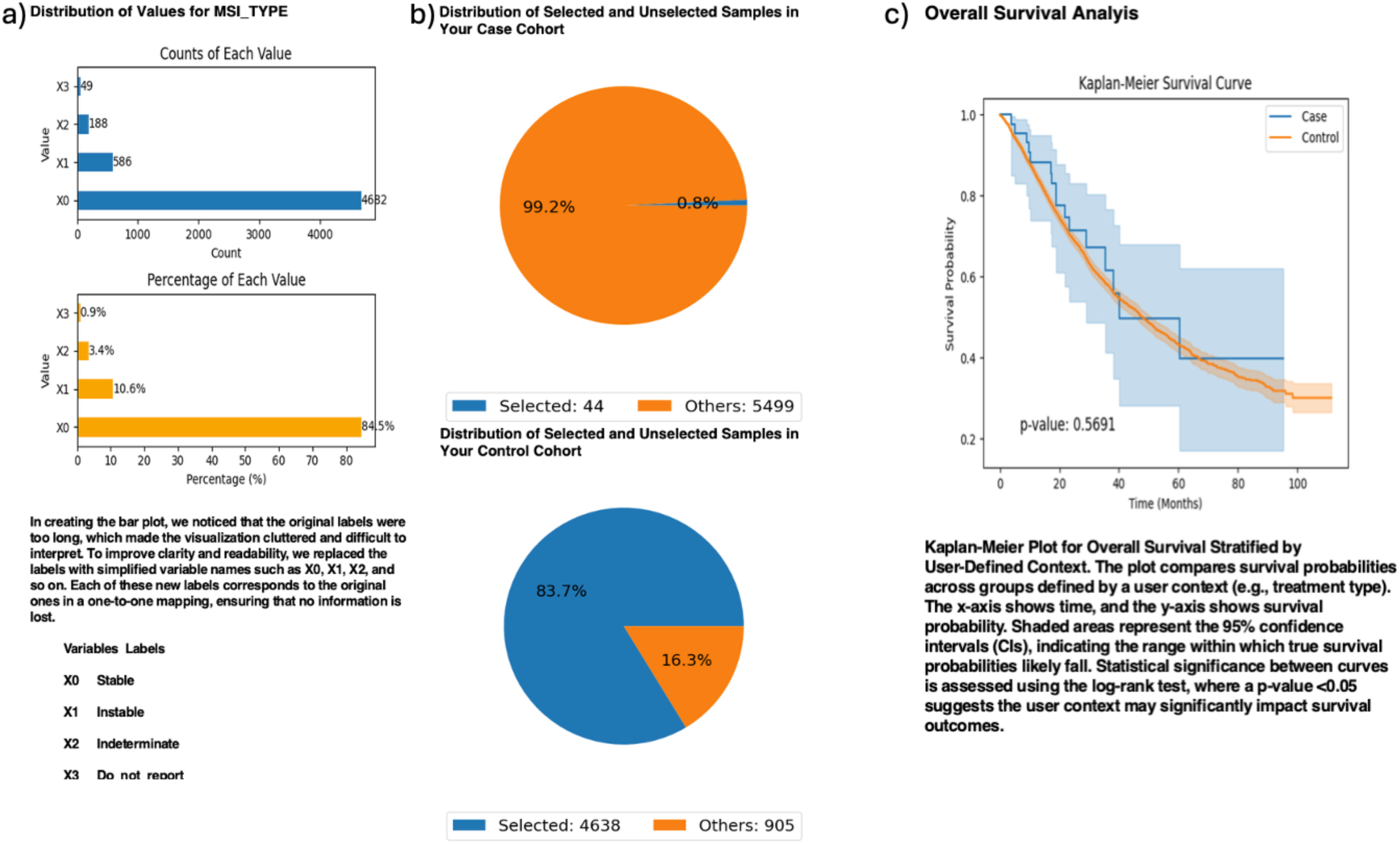
AI-HOPE-JAK-STAT analysis of JAK1 mutation status in microsatellite stable colorectal cancer. This figure illustrates the use of AI-HOPE-JAK-STAT to investigate overall survival differences in colorectal cancer (CRC) patients with stable microsatellite status (MSI-Stable), stratified by JAK1 mutation status. The case cohort includes MSI-Stable patients harboring JAK1 mutations (n = 44), while the control cohort includes MSI-Stable patients without JAK1 mutations (n = 4,638). a) Bar plots display the distribution of MSI types across the full dataset using simplified labels (e.g., X0 = Stable, X1 = Instable). MSI-Stable tumors represent the majority of samples, confirming sufficient data to evaluate JAK1-specific outcomes within this molecular context. b) Pie charts show the relative proportion of selected samples in the case and control cohorts. The JAK1-mutant group comprises only 0.8% of the dataset, compared to 83.7% for the JAK1 wild-type group within MSI-Stable cases, underscoring the rarity of this alteration. c) Kaplan-Meier survival analysis reveals no statistically significant difference in overall survival between the two cohorts (p = 0.5691). Although the JAK1-mutant group shows a slightly different survival trajectory, the wide confidence intervals and overlapping curves indicate no clear association with survival in this subset. These results suggest that JAK1 mutation status may not independently influence prognosis among MSI-Stable CRC patients and highlight the importance of considering broader molecular contexts in precision oncology studies.

Further, we analyzed STAT5B mutation status in primary tumor samples. Patients with STAT5B-mutated primary tumors (n = 109) experienced significantly better overall survival than those without such alterations (n = 3,945; p < 0.0001; Fig. S2). Odds ratio testing revealed that STAT5B-mutated tumors were more frequently located in the colon than in other sites (OR = 1.949, p = 0.002), suggesting anatomical preferences in mutation distribution and potential therapeutic relevance.

Finally, we explored stage-specific survival patterns among JAK3-mutated CRC patients. When stratified by disease stage, individuals diagnosed at Stage I–III (n = 142) exhibited significantly better outcomes than those with Stage IV tumors (n = 36), with highly significant survival differences (p < 0.0000; Fig. S3). These findings underscore the strong prognostic impact of clinical stage even within molecularly defined subgroups.

Together, these analyses establish AI-HOPE-JAK-STAT as a robust, interactive platform capable of uncovering clinically relevant associations between JAK/STAT signaling alterations and survival outcomes across stratified CRC cohorts. By unifying natural language interfaces with integrated bioinformatics, the system supports reproducible, real-time analysis of complex cancer datasets—offering new pathways toward individualized treatment strategies.

## Discussion

This study presents AI-HOPE-JAK-STAT, a novel conversational Artificial Intelligence platform designed to interrogate the clinical-genomic landscape of JAK/STAT signaling alterations in CRC. By combining a natural language interface with automated, reproducible bioinformatics workflows, the system addresses longstanding limitations in accessibility and flexibility of pathway-specific data analysis—particularly for EOCRC and other clinically stratified subgroups.

Unlike conventional tools that rely on manual curation or scripting expertise, AI-HOPE-JAK-STAT enables users to conduct complex, multi-layered analyses through natural language queries. Across validation and exploratory tasks, the platform replicated known associations—such as the prognostic impact of JAK/STAT alterations in EOCRC—and uncovered novel trends in treatment response and mutational co-enrichment. In doing so, AI-HOPE-JAK-STAT has demonstrated that conversational AI can serve as a scalable and robust solution for precision oncology research.

Our findings reinforce the biological and clinical relevance of JAK/STAT signaling in CRC. In early-onset patients of both Hispanic/Latino (H/L) and NHW backgrounds, alterations in pathway genes such as JAK1, JAK3, and STAT5B were associated with improved survival—particularly among those receiving FOLFOX chemotherapy. These results align with preclinical literature suggesting that JAK/STAT dysregulation may sensitize tumors to immune-mediated or cytotoxic interventions. Moreover, the observed stage-specific survival differences among JAK3-mutant tumors (Stage I–III vs. Stage IV) suggest that clinical context significantly modulates the prognostic impact of these alterations.

Beyond outcome comparisons, AI-HOPE-JAK-STAT was also effective in evaluating population-level enrichment. For example, STAT5B mutations were found to be significantly more prevalent in colon versus rectal tumors, an anatomical preference that may reflect divergent etiologies or tissue-specific gene regulation. Conversely, JAK1 mutations within microsatellite-stable (MSS) tumors did not exhibit a survival effect, emphasizing the importance of molecular context in interpreting prognostic significance. Such nuanced insights are rarely obtainable without substantial analytic infrastructure, yet were generated in seconds using AI-HOPE-JAK-STAT’s conversational engine.

A key strength of the platform lies in its adaptability across user-defined comparisons— age, ancestry, tumor stage, and treatment regimen—all seamlessly integrated through structured outputs. This flexibility positions AI-HOPE-JAK-STAT as not just an analytics tool, but a collaborative system for hypothesis testing, education, and translational discovery. The integration of retrieval-augmented generation (RAG) further enhances interpretability by contextualizing results within the existing literature, supporting evidence-based insight generation.

Nevertheless, several limitations warrant discussion. The reliance on public datasets may restrict generalizability due to variable sample representation. Although AI-HOPE-JAK-STAT supports stratification by ancestry and tumor type, broader deployment will benefit from inclusion of additional multi-omic layers (e.g., RNA-seq [10,39], spatial biology data [40–42]) and prospective clinical datasets. Additionally, while the natural language interface promotes usability, performance can vary based on query specificity; future iterations may incorporate query refinement features and dialogue memory to improve user interaction.

Despite these limitations, the platform’s performance in recapitulating prior findings, identifying novel associations, and supporting pathway-level analyses without coding expertise reflects a meaningful advancement in AI-enabled cancer research. AI-HOPE-JAK-STAT not only complements traditional tools like cBioPortal and Xena but offers unique advantages in speed, flexibility, and hypothesis exploration.

## Conclusion

AI-HOPE-JAK-STAT represents a significant innovation in precision oncology infrastructure, enabling natural language-guided integration of genomic and clinical data to decode the role of JAK/STAT pathway alterations in CRC. By facilitating real-time survival analysis, mutation profiling, and population-level stratification, the platform empowers researchers and clinicians to extract actionable insights from complex datasets. As CRC research moves toward individualized therapies and immunologic profiling, tools like AI-HOPE-JAK-STAT will be instrumental in accelerating biomarker discovery and supporting context-aware clinical decision-making.

## Data Availability

All data used in the present study is publicly available at https://www.cbioportal.org/ and https://genie.cbioportal.org. Additional data can be provided upon reasonable request to the authors.

---

**Figure S1.**
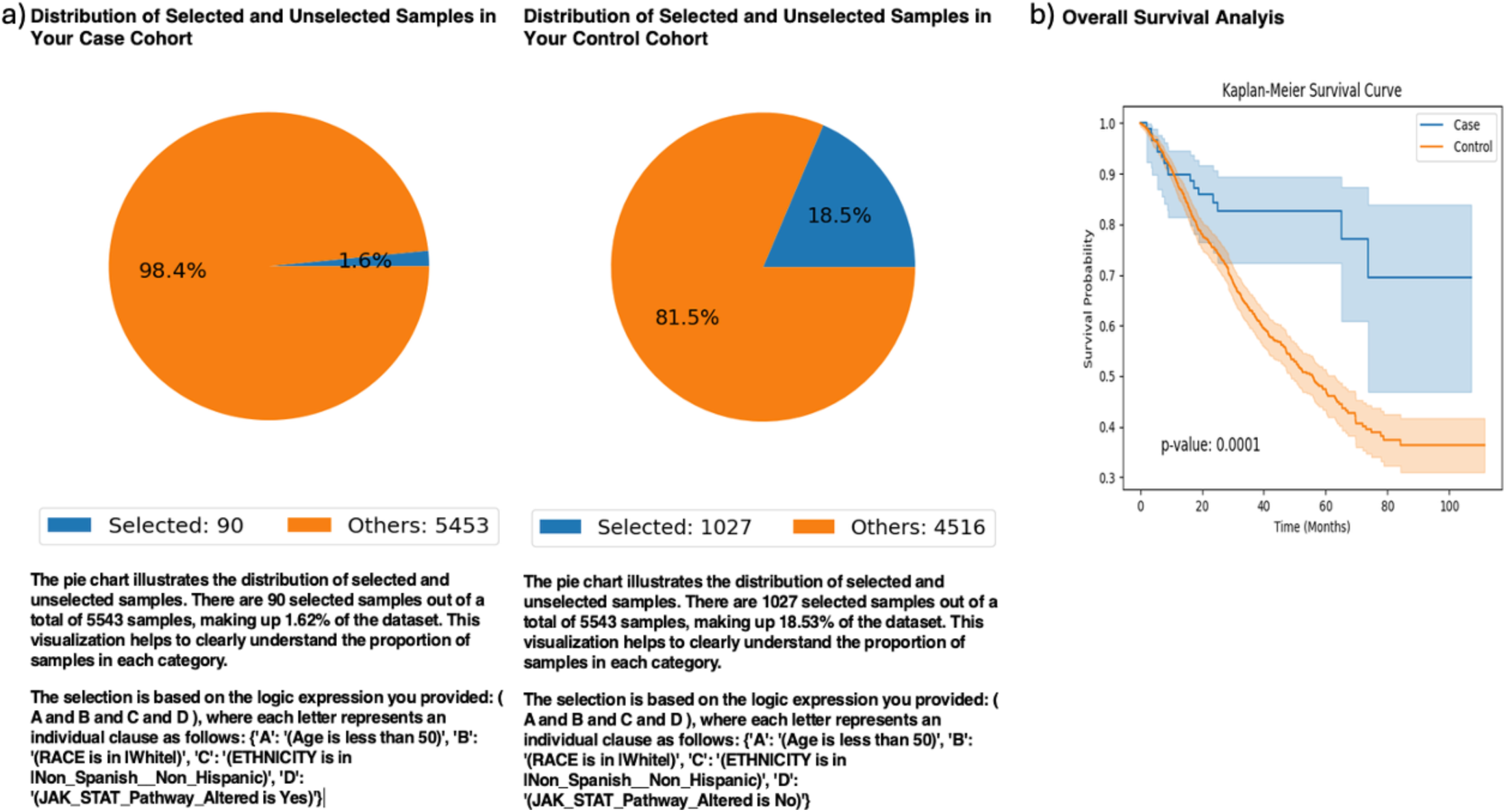
AI-HOPE-JAK-STAT analysis of early-onset colorectal cancer in patients stratified by JAK/STAT pathway alteration status. This figure presents the outcome of a natural language-driven query performed using AI-HOPE-JAK-STAT to evaluate the association between JAK/STAT pathway alterations and overall survival in early-onset colorectal cancer (EOCRC) patients of Non-Hispanic White (NHW) background. a) Pie charts display the proportion of selected versus unselected samples within each cohort relative to the full colorectal cancer dataset (n = 5543). The representation highlights the relative infrequency of JAK/STAT pathway mutations in this subgroup, yet sufficient for comparative survival analysis. b) The Kaplan-Meier survival plot demonstrates a marked and statistically significant difference in survival outcomes between the two groups. Patients with JAK/STAT alterations (blue) exhibited considerably improved survival compared to their non-altered counterparts (orange). The p-value of 0.0001 from the log-rank test confirms the robustness of this association. The narrow confidence intervals further support the reliability of the observed survival benefit, suggesting a potentially protective or biologically meaningful role of JAK/STAT pathway alterations in EOCRC among NHW individuals.

**Figure S2.**
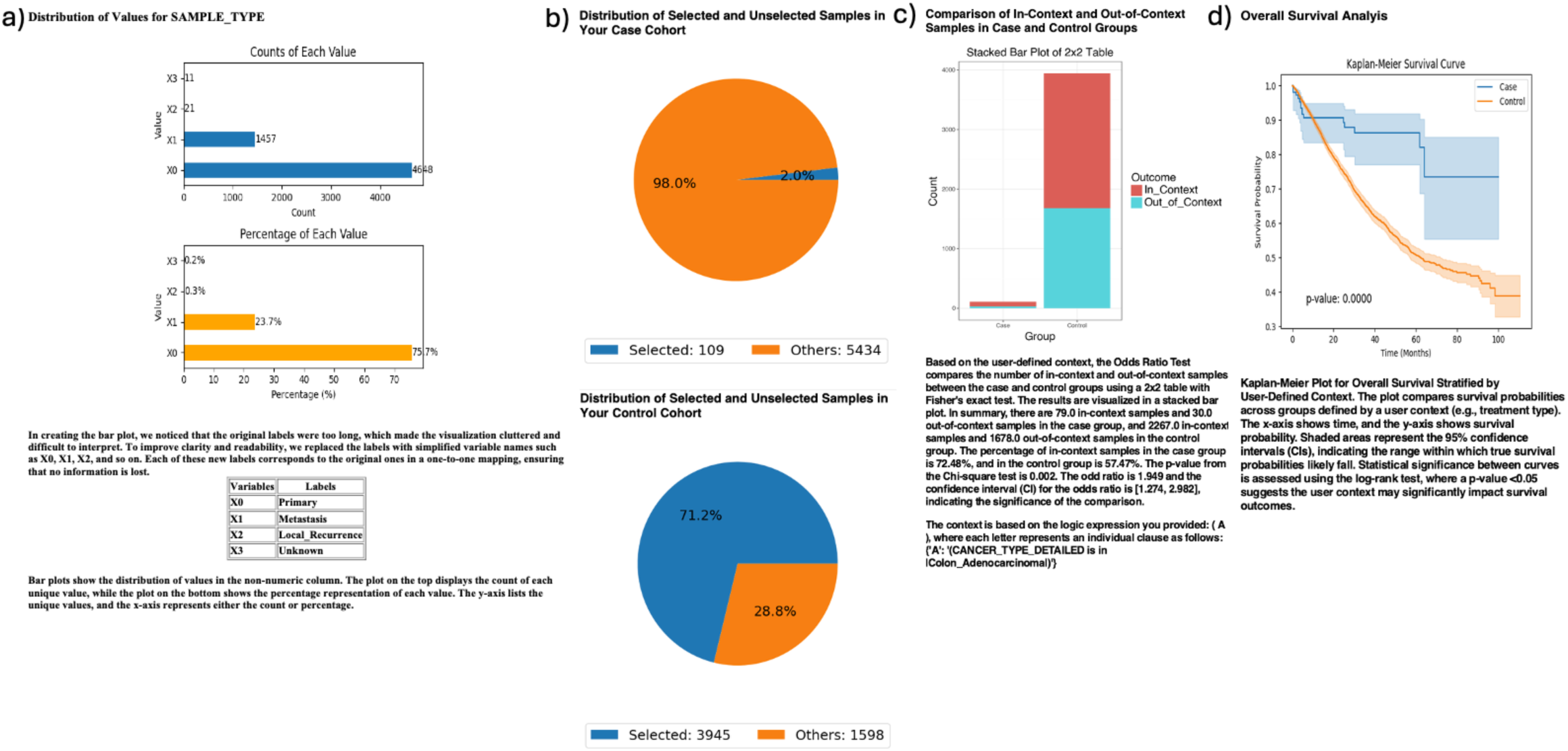
AI-HOPE-JAK-STAT analysis of STAT5B-mutant primary colorectal tumors with odds ratio context in colon adenocarcinoma. This figure highlights the application of AI-HOPE-JAK-STAT to assess survival outcomes and mutation enrichment among colorectal cancer (CRC) patients with STAT5B mutations in primary tumor samples. The case cohort includes primary tumors harboring STAT5B mutations (n = 109), while the control cohort includes primary tumors without such mutations (n = 3,945). a) Bar plots summarize the distribution of sample types across the dataset using simplified labels (e.g., X0 = Primary, X1 = Metastasis), confirming that the majority of samples are primary tumors. b) Pie charts illustrate the selection of case and control cohorts based on mutation status. STAT5B-mutated primary tumors represent 2.0% of the dataset, while STAT5B wild-type cases account for 71.2%, indicating sufficient data availability for comparative analysis. c) An odds ratio test evaluates the enrichment of colon adenocarcinoma cases across the defined cohorts. A 2×2 table compares in-context (colon adenocarcinoma) and out-of-context samples in both case and control groups. Among the case group, 72.48% were in-context, compared to 57.47% in the control group. The resulting odds ratio is 1.949 (95% CI: 1.274–2.982; p = 0.002), suggesting a statistically significant association between STAT5B mutations and colon tumor location. d) Kaplan-Meier survival analysis demonstrates significantly better overall survival in patients with STAT5B mutations (blue) compared to those without (orange), with a p-value of < 0.0001. The clear separation of survival curves and narrow confidence intervals support a potential prognostic benefit of STAT5B mutation status in CRC patients with primary tumors.

**Figure S3.**
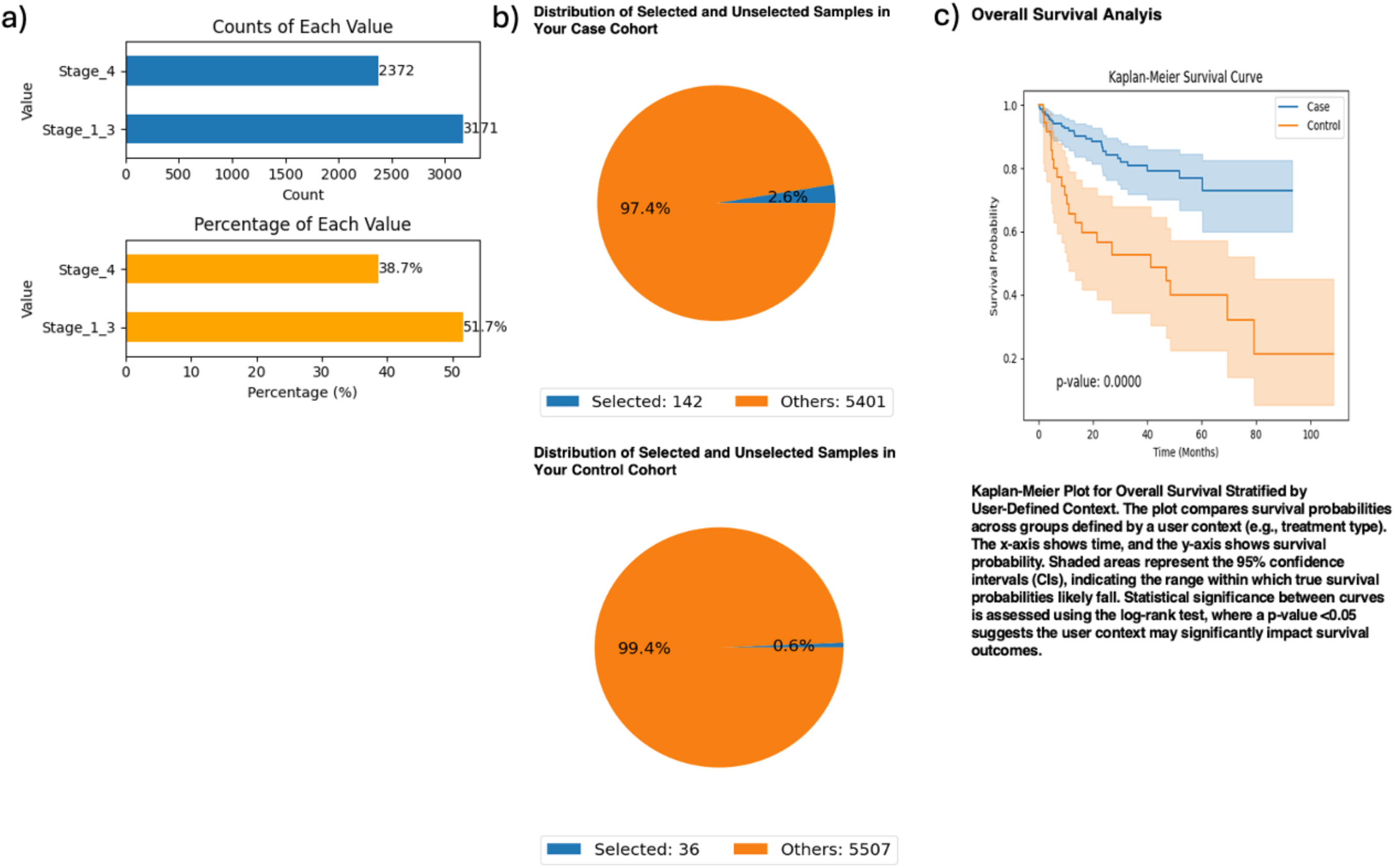
AI-HOPE-JAK-STAT analysis of colorectal cancer patients with JAK3 mutations stratified by tumor stage. This figure demonstrates how AI-HOPE-JAK-STAT enables stratified survival analysis in colorectal cancer (CRC) patients carrying JAK3 mutations, comparing those with early-stage (Stage I–III) versus advanced-stage (Stage IV) disease. a) Bar plots show the distribution of patients across tumor stages, with 51.7% in Stage I–III (n = 3,171) and 38.7% in Stage IV (n = 2,372). These distributions confirm the presence of adequate sample sizes across stage groups to support comparative analyses. b) Pie charts display the relative proportions of selected JAK3-mutant patients in each cohort. The early-stage group includes 142 samples (2.6% of the total dataset), while the advanced-stage group includes 36 samples (0.6%). This visualization reflects the lower prevalence of JAK3 mutations among late-stage CRC cases. c) Kaplan-Meier survival analysis reveals a highly significant difference in overall survival between the two groups (p < 0.00001). Patients with JAK3 mutations diagnosed at Stage I–III (blue) demonstrate markedly improved survival relative to those diagnosed at Stage IV (orange). The wide separation between survival curves and narrow confidence intervals reinforce the strength of the association, suggesting that tumor stage remains a strong prognostic factor even among JAK3-mutated CRC patients.

